# Comprehensive and Temporal Surveillance of SARS-CoV-2 in Urban Water Bodies: Early Signal of Second Wave Onset

**DOI:** 10.1101/2021.05.08.21256881

**Authors:** Manupati Hemalatha, Athmakuri Tharak, Harishankar Kopperi, Uday Kiran, C. G. Gokulan, Rakesh K Mishra, S Venkata Mohan

## Abstract

The possible faecal-oral transmission of SARS-CoV-2 through domestic discharges has emerged as a serious public health concern. Based on persistence of the virus in environment, the wastewater-based epidemiology (WBE) enabled the surveillance of infection in a community. The water bodies connected to the anthropogenic activities have strong possibility of presence of the SARS-CoV-2 genetic material. In this work, we monitored urban, peri-urban and rural lakes in and around Hyderabad as a long-term surveillance study for presence of enteric virus SARS-CoV-2 gene fragments. The study time of seven months coincided with the first and second wave of COVID-19 infection. The study depicted differential viral RNA copies in the urban lake with high viral load observed during the peaks of wave I and wave II. Distinct variability in viral genes detection was observed amongst all five lakes which were in concordance with the human activity of the catchment area. The SARS-CoV-2 genes were not detected in peri-urban and rural lakes, whereas the urban lakes having direct functional attributes from domestic activity, in the community showed presence of viral load. The outcome of the study clearly shows that the urban water streams linked with domestic discharge will function as a proxy for wastewater epidemiological studies. The surge in viral gene load from February 2021 sample suggests the on shoot of the second wave of infection, which correlated well with the prevailing pandemic situation. Implementation of regular WBE based monitoring system for the water bodies/wastewater in the urban and semi-urban areas will help to understand the outbreak and spread of virus in the community.

## 1. Introduction

The novel corona virus SARS-CoV-2 challenged the existing diagnostic and clinical management. The transmission of the viral genetic material via faecal-oral route, wastewater-based epidemiology (WBE) studies gained phenomenal attention (Ahmed et al., 2020; Daughton et al., 2018; Mao et al., 2020; Venkata Mohan et al., 2021; Wu et al., 2020). The WBE studies offer unbiased infection status and dynamics in a given population and also provide early warning for better management of infection (Hemalatha et al., 2021; Kopperi et al., 2021; Sims and Kasprzyk-Hordern 2020; Tharak et al., 2021; Zahedi et al., 2021). The occurrence and detection of different pathogenic viruses such as polio, SARS and MERS in wastewater has been shown previously (Tran et al., 2020; Drosten et al., 2013). Unlike polio virus the transmission of SARS-CoV-2 through water contamination is not well established, only one report showed successful culturing of SARS-CoV-2 virus from wastewater (Randazzo et al., 2020). The independent previous studies showed presence and persistence of SARS-CoV-2 in sewage water and water bodies such as lakes and rivers, because of which the possible faecal-oral transmission cannot be neglected (Guerrero-Latorre et al., 2020; Annalaura et al., 2020). It is largely agreed that that different environmental factors like temperature, presence of different chemical contaminants and detergents play detrimental role on stability of virus/viral particles in sewage water (Mattle and Kohn, 2012; Sigstam et al., 2013; Venkata Mohan et al., 2021). The possibility of viral contaminated water transmission cannot be overlooked in light of all these factors in water bodies. Considering the significant human activities near water systems, it is important to conduct the long-term surveillance for possible viral contamination in water systems (Guerrero-Latorre et al., 2020). Urban water bodies function as a proxy to the anthropogenic activities surrounding it (Venkata Mohan et al., 2009:(Kumar et al., 2016). Despite the viral genetic material/fragments in sewage or in water bodies are non-viable and does not infect the community, however, can be used as surveillance tool to understand the infection onset and spread. In this work we monitored an urban lake (UL-1) for seven months for the detection SARS-CoV-2 virus material along with less polluted urban lakes (UL-2 and UL-3), peri-urban (PL) and rural lake (RL).

## 2. Materials and Methods

### 2.1. Sampling Sites and Collection

Water samples from selected lentic water bodies viz., urban, peri-urban and rural areas in and around Hyderabad, were collected by employing grab sampling protocol between 8:00 to 10:00 am. The samples were collected on the days, when there was no rainfall event in last 48 hours. Sterile sampling bottles were used for the sampling, with 20 mL 0.1% disinfectant (0.1% Sodium Hypochlorite) (Kopperi et al., 2021; Hemalatha et al., 2021). Three lakes in the urban zones of Hyderabad viz., Peddacheruvu/Nacharam lake (UL-1, 17.42°N 78.55°E), Hussain Sagar lake (UL-2, 17.41°N, 78.47°E) and Nizam Talab (also called Turkha Cheruvu) lake (UL-3, 17.52°N, 78.38°E) were chosen for the study (Fig 1). Edulabad lake (17.42°N, 78.69°E) (PL) and Potharaju lake (17.40°N, 78.71°E) (RL) were sampled as referral lakes under per-urban and rural functions respectively. Long term surveillance (weekly and monthly) was performed for UL-1 with two samples (lake and lake outlet) for four weeks (Week-1 (7/10/2020); Week-4 (28/10/2020); Week-5 (04/11/2020)); Week-6 (18/11/2020)) and monthly monitoring for 7 months (October 2020 to April 2021) were collected. Week 2 (14/10/2020) and Week 3 (21/10/2020) samples were not collected due to the occurrence of multiple rainfall events.

**Fig. 1:**
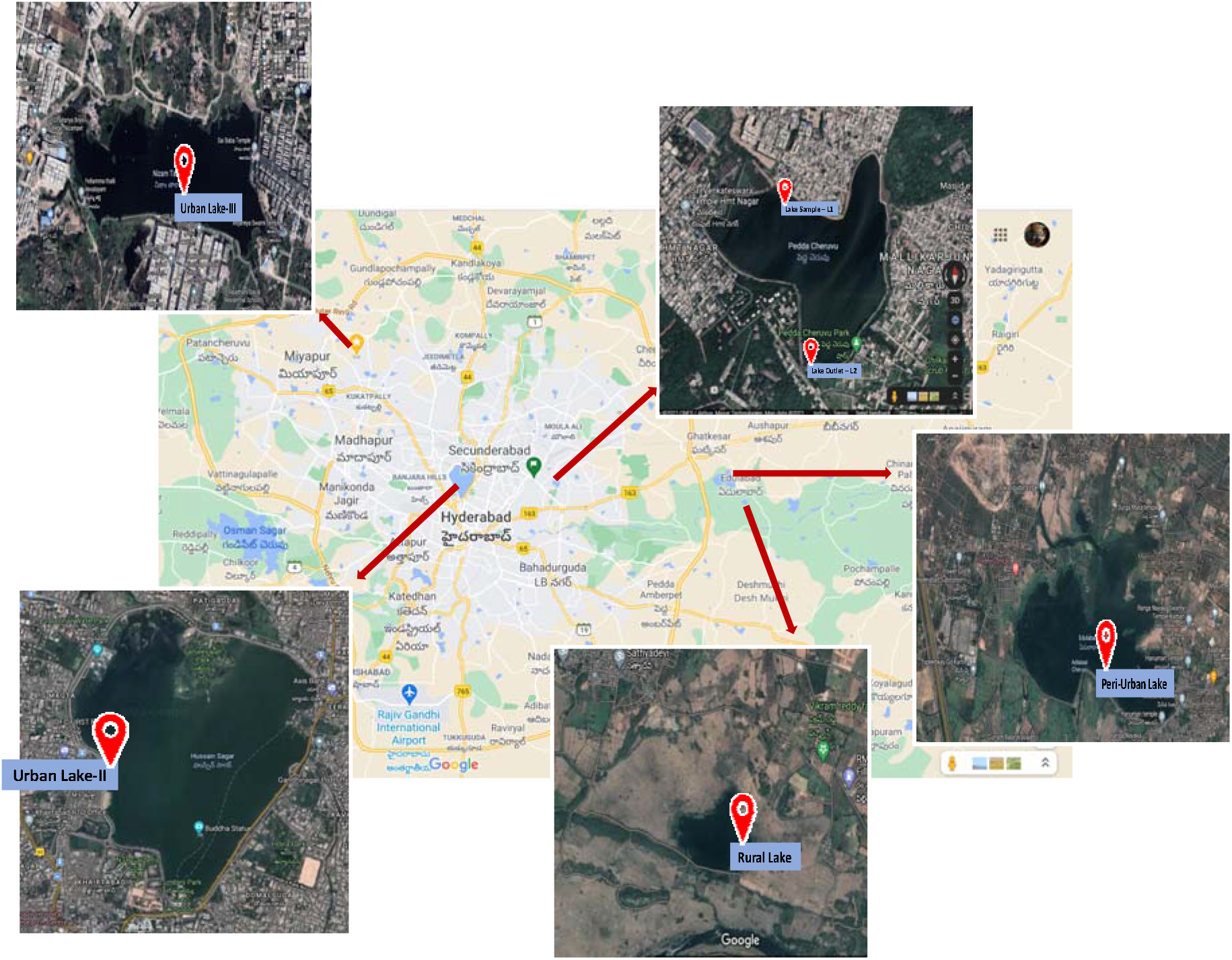
Map illustrating the point of sample collection with reference to lakes in and around Hyderabad (Courtesy: Google Map)

### 2.2. Sample Fractionation

After collection, samples were packed in a disposable pack to avoid leakage during the transportation. Samples were brought to lab within 3 h of sample collection and processed within 24 h. Samples were initially filtered using 1 mm filter paper to remove larger debris followed by 0.2 μm filtration to separate suspended solids. A 60 mL of filtrate was further subjected to ultrafiltration (4000 rpm; 4 °C; 10 min) using 15 mL Amicon (30 kDa Amicon® Ultra-15 (Merck Millipore)) to concentrate to 600 μL.

### 2.3. Nucleic Acid Extraction from Concentrate Fraction and RT-PCR

A 150 μL of concentrate was used for RNA extraction using a viral RNA isolation kit (QIAamp, Qiagen) as per manufactures protocol. Sterile material devoid of DNA/RNA cross-contamination and RNase-free water was used for RNA isolation. Isolated RNA was quantified by RT-PCR Detection Kit (Shanghai Fosun Long March Medical Science Co., Ltd, China) approved by FDA (Food and Drug Administration, USA). Fosun RT-PCR contains primers, probes (chromophore labelled) encoding for envelope protein-coding gene (E-gene; ROX), nucleocapsid gene (N-gene; JOE), and open reading frame1ab (ORF1ab; FAM) of SARS-CoV-2. The RT-PCR reaction for SARS-CoV-2 detection includes Reverse transcription for 15 min at 50°C and the Initial denaturation for 3 min at 95°C for 3 minutes followed by 45 cycles at 95°C for 5 seconds and 60°C for 40 seconds. Signals from the chromophore probes ROX (E gene), JOE (N gene), FAM (ORF1ab), and CY5 (Internal reference) were collected by the fluorescence channels at 60°C. All the amplifications include positive and negative controls provided in the fosun RT-PCR kit. The negative controls were clean and C_T_ values for positive samples were in a match with the manufacturer’s recommendation. All the reactions were performed in triplicates in a Biosafety level 2 (BSL-2) lab.

### 2.4. Statistical Analysis

From the C_T_ value obtained RNA copies/L of water using the linear fit equation of E-gene (Eq. 1) and relative standard deviation (RSD) (Eq. 2) for the individual gene were calculated (Hemalatha et al., 2021). RNA copies/L of wastewater was calculated using the linear fit equation of the E-gene (Hemalatha et al., 2021). Relative standard deviation (RSD) was calculated for the C_T_ value of individual genes based on the Eq. 4, where, 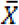 is the mean of C_T_ value and ‘S’ is the standard deviation (Hemalatha et al., 2021).

## 3. Results and Discussion

### 3.1 Surveillance of Targeted Gene Fragments -Urban Lake-I

Peddacheruvu-Nacharam Lake (UL-1) selected for long term viral RNA surveillance is a manmade lake spreading over 90 acres surround by a population of ∼10,00,000. The catchment area of water body is surrounded by densely populated anthropogenic activities and receives majorly three-point sources viz., treated discharge of 10 MLD capacity sewage treatment plant (STP) and the other two domestic discharges with relatively low flow. The lake water samples analysed showed COD of 152 mg/L with TDS of 800 mg/L and TSS of 110 mg/L respectively. The unfiltered sample of lake showed a yellowish tinge while the lake appears dark grey in colour with eutrophication behaviour.

#### 3.1.1 Weekly Monitoring

A total of 8 samples over a period of six weeks were collected (07/10/2020 (week 1); 28/10/2020 (week 4); 04/11/2020 (week 5); 18/11/2020 (week 6)) accounting 4 samples each from Peddacheruvu lake (UL1) and its outlet point. Week 2 (14/10/2020) and Week 3 21/10/2020) were not collected due to the occurrence of multiple rainfall events. All the lake samples showed positive signal for the target genes (E gene, N gene and ORF1ab). The C_T_ value of E-gene in lake samples varied between 28.16±0.46% and 23.04±12.02% with continuous decrement from week 1 (28.16±0.46%), Week 4 (24.37±1.23%), Week 5 (24.81±1.70%) and week 6 (23.04±3.86%) with an average of 25.10±3.85%. A similar trend was observed in both N-gene (week 1: 27.04±0.81%; week 4: 22.35±0.62%; week 5: 23.32±1.83%; week 6: 22.27±1.54%) and ORF1ab (week 1: 27.73±0.75%; week 4: 26.62±0.80%; week 5: 25.69±1.86%; week 6: 22.23±2.64%) with an average of 23.74±3.7% and 25.56±3.40% respectively. The continuous decrement in C_T_ of all the targeted genes represents the increase in viral load in the community surrounding the UL-I with time.

Viral RNA copies were calculated based on the C_T_ of E-gene using linear fit equation (Hemalatha et al., 2021). The increase in viral RNA copies/L with respect to time was observed for lake samples which ranged between 26,927 to 9,75,668. The highest RNA copies/L was observed in Week 6 (9,75,668) followed by Week 5 (2,82,039), Week 4 (3,83,969) and Week 1 (26,927 RNA copies/L) (Fig. 2) In spite of the overall decrement with time, the marginal increase in C_T_ values (∼0.5) with reference to E-gene between Week 4 to week 5 resulted in decrement of RNA copies significantly. The range of RNA copies correlated well with the infection spread of the community near the UL-I (Tharak et al., 2021).

**Fig. 2:**
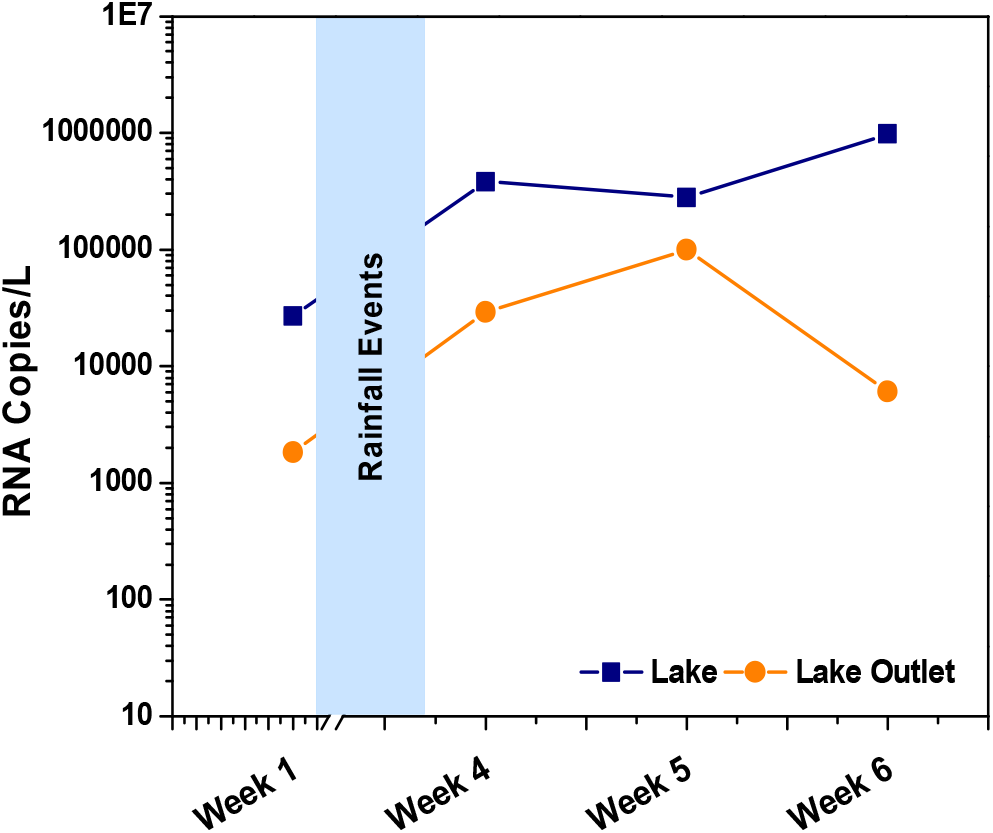
RNA copies calculated based on linear fit equation of E-gene of samples collected weekly from lake and lake outlet samples of UL-1.

Interestingly 75% of Lake Outlet samples showed positive signals for the viral genetic material presence. E-genes was detected in all the outlet samples, while N-gene was detected in Week 1, 4, and 5 but ORF1 gene was detected in week 1 and 2 samples only. The trend of C_T_ values showed relatively lower viral load in outlets samples when compared to lake samples. Comparatively lower RNA copies (1,823/L) were observed in Week 1 while Week 5 depicted higher RNA copies (99,914/L) (Fig. 2).

#### 3.1.2 Monthly Monitoring

A total of 16 samples were collected from October 2020 to April 2021. As there is a rise in COVID-19 positive cases from March 2021, the sampling was performed twice in April 2021 (01/04/2021 and 15/04/2021 (mid-April)). All the lake samples detected all the three genes. The E-gene C_T_ varied significantly between 31.94±2.15% and 23.04±12.02% and correlates well with the First and Second Wave scenario. A C_T_ of 28.16±0.46% was detected for October 2020 sample and further decreased to 24.81±1.70% and 23.04±12.02% for November 2020 and December 2020 respectively. A sharp rise in C_T_ values was observed for January 2021 and February 2021 samples which was 31.94±2.15% and 31.34±1.97% respectively with a slight increase of ∼0.6 C_T_. From February 2021 a marked decrement in C_T_ was noticed followed by March 2021 and April 2021 samples (28.75±0.54% and 27.38±2.00%) representing the onset of second wave. A similar trend was observed for the other two targeted genes. The samples from October 2020 to December 2020 showed a decrease in C_T_ values for N-gene from 27.04±0.81% (Oct 2020), 23.32±1.83% (Nov 2020) and 23.32±1.83% (Dec 2020) followed by increment in January 2021 (30.10±1.62%). A further decrease in N-gene was observed from February 2021 to mid-April 2021 (Feb 2021 (29.04±1.58%); March 2021 (28.47±0.09%); 01 April 2021 (27.11±0.62%); 15^th^ April 2021 (26.03±2.09%)). For ORF1ab, the decrease in C_T_ was observed from October 2020 (27.73±0.75%) followed by November 2020 (25.69±1.86%) and December 2020 (22.23±10.21%). No detection of ORF1ab was observed in January 2021 samples. Whereas, February 2021 sample showed detection (30.53±5.54%) followed by persistent decrement (26.46±2.62% (March 2021); 25.26±1.81% (01 April 2021)). An increase in ∼2 C_T_ was observed from 01 April 2021 to mid-April 2020 which showed the C_T_ of 27.13±2.17% in lake samples. The viral load gets affected by the lake ecosystem condition as well as the quantity of domestic/wastewater discharge and their quality apart from climatic conditions prevailing (Westhaus et al., 2021). The outlet of UL-1 showed positive to two genes (E-gene and N-gene) among the three targeted genes. The detection was observed from October 2020 to February 2021 whereas, March 2021 and April 2021 samples showed no detection for targeted genes. The decrease in E-gene C_T_ was observed from 32±0.89% (October 2020) to 26.29±1.40% (November 2020) which later showed an increasing trend till February 2021 (32.32±0.09%). Similarly, N-gene C_T_ was observed to decrease from 32.43±1.26% (October 2020) to 24.89±3.37% (November 2020) which later showed an increasing trend till February 2021 (32.26±0.06%) with no detection of ORF1ab in outlet samples.

Viral RNA copies were calculated for the monthly monitoring samples using E-gene linear fit Equation (Hemalatha et al., 2021) where the higher RNA Copies of 9,75,668/L for December 2020 representing peak infection during end of first phase. A lower copy number was observed in January 2021 (3,128 RNA copies/L), February 2021 (2,881 RNA copies/) followed by a slight increase in March 2021 (4,665 RNA copies/L) (Fig. 3). Further April 2021 samples showed an increase in trend of RNA copies from 17,804 RNA copies/L (01 April 2021) and 46,217 RNA copies/L (Mid-April 2021) (Fig. 3) depicting the emergence of second wave. The infection rate is more or less equal to the COVID-19 pandemic Wave-I. RNA copies of lake outlet samples were also followed the same trend as highest copies were observed in November 2020 (99.914 RNA copies/L) followed by December 2020 (6,090 RNA copies/L) (Fig. 3).

**Fig. 3:**
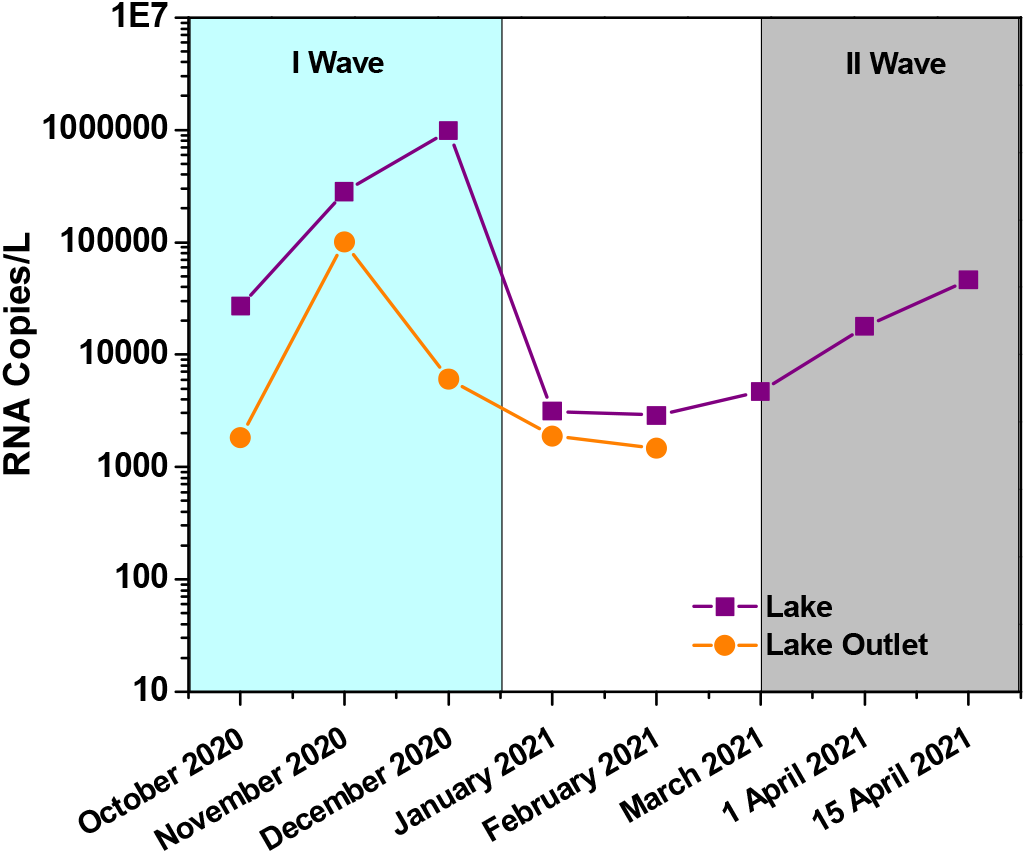
RNA copies calculated based on linear fit equation of E-gene of samples collected monthly from lake and Lake Outlet of UL-1.

### 3.2 Surveillance of Targeted Gene Fragments-Urban Lake-2

Nizam Talab Lake ((UL-2) located in Pragathi Nagar area of Kukatpally, Hyderabad covers nearly about 35 acres with a residing population of ∼1,50,000 on the catchment area. UL-2 receives three-point sources (domestic discharges) and has one outlet. The lake showed COD of 134 mg/L and TDS of 524 mg/L. Visually the lake was transparent in color without any sign of eutrophication. The samples collected for three months, from the end of COVID-19 Wave-1 till the onset and persistent phase of COVID-19 Wave-2 (December 2020, January 2021 and February 2021). UL-2 samples showed the presence of N-gene and ORF1ab. The decrease in trend of C_T_ for N-gene and ORF1ab was observed. N-gene decreased from 33.62±1.33% (Dec 2020) followed by Jan 2021 (30.02±1.69%) and Feb 2021 (29.08±1.65%). Whereas ORF1ab C_T_ values sowed marginal decrement from December 2020 to January 2021. RNA copies were not calculated as the sample showed no detection of E-gene for all the samples. The lake outlet samples did not detected for the three targeted genes. Relatively young lake, non-signs of eutrophication and self-regeneration capacity might be the probable reasons for not detection all the genes in the lake water samples even though untreated domestic sewage discharge was there. Also, the composition of domestic discharges specifically with surfactants will also influence the viral genetic material presence in lake water body.

### 3.3 Surveillance of Targeted Gene Fragments-Urban Lake-3

Hussain Sagar Lake (UL-3), a heart of Hyderabad is an important man-made lake with a spread of 1409 acres and depth of 32 feet. This lake has majorly point sources (treated sewage) of 50 MLD from two STPs. The COD of lake showed 225 mg/L with TDS and TSS of 720 mg/L and 125 mg/L respectively. The presence of aquatic microflora was evident with signs of eutrophication. In the case of Husain Sagar (UL-3), the samples were collected during the two infection peaks (waves) phases of COVID-19 (July 2020 and April-2021). Both the samples showed the amplification of two (E-gene and N-gene). The detected samples showed higher E-gene with C_T_ of 30.82±0.06% and 33.44±0.12% for July 2020 and April 2021 respectively. Whereas the N-gene C_T_ of 31.18±0.11% (July 2020) and 31.96±0.16% (April 2021) was observed. RNA copies of 4,160/L and 664 RNA copies/L were observed in July 2020 and April 2021 respectively. This might be possible as the lake receives majority of treated water as discharge and also due to dilution effect apart from the ecosystem nature.

### 3.4 Surveillance of Targeted Gene Fragments-Peri-urban/Rural Water Bodies

The Peri-urban lake (Edulabad Lake, PL) is located near to Ghatkesar covers about 1236 acres of area. The lake catchment area is covered by villages and agricultural fields. It gets both non-point and point sources accounting for domestic discharges and agricultural runoff. The physico-chemical characteristic of the PL was with 176 mg/L COD, 443 mg/L TDS and 63 mg/L of TSS. The rural lake named Pothuraju lake (RL) which is located in south-east direction of PL is surrounded by agriculture fields and majorly receives non-point runoff showed to have COD of 127 mg/L, TDS of 337 mg/L and TSS of 46 mg/L of TSS. Samples from both PL and RL were collected during second wave phase (01/04/2021). Both these lakes were used as a referral water bodies for comparison purpose. Samples from PL and RL were negative for the targeted genes which might be because of the nature of lakes.

### 3.5 Comparative analysis

A comprehensive long-term monitoring of different types of lentic water bodies located in urban, semi-urban and rural locations showed the presence of RNA genetic material of virus attributing to the associated functional activities of the lake catchment area. Urban lakes encompassing the domestic activities showed the prevalence of viral load and may be consider as proxy for WBE studies to assess the community infection rate. The receiving of domestic discharge from the population residing around the catchment area forms a major basis for this. UL-1 samples showed positive signal for all the three genes, UL-2 and UL-3 showed positive signal for two genes out of three (UL-2 (N-gene and ORF1ab) and UL-3 (E-gene and N-gene)). The referral lakes PL and RL did not show detection for the targeted genes. From the results it is clear that urban lakes are impacted probably due to non-treated sewage discharge resulted in high viral load. The trend of RNA copies curve (with reference to UL-1) is correlating with the dynamics of COVID cases matching with Wave I and Wave II scenario in India. This observation indicates a clear-cut function of the water bodies to act as a proxy for surveillance studies to predict the epidemic/pandemics as early warning signal (as part of WBE studies), to assess the infection rates during the ongoing pandemic and to understand the dynamics of viral load pertains to the community in the catchment area.

## 4. Conclusions

The SARS-CoV-2 gene fragments have been detected were clearly in the urban lakes which are surrounded by the anthropogenic activities. Also, the dynamic of viral load showed the proximity to understand the infection rates as well as also served as early warning signal. The surge in February 2021 sample showed the on shoot of the second wave of infection which correlated well with the prevailing pandemic situation. The reference water bodies (peri-urban and rural lakes) did not detected SARS-CoV-2 fragments. The functional attributes of lake to anthropogenic activity will be the guiding factor for the viral presence. The presence of viral fragments and its load can be used as surveillance tool to understand the infection spread. This study depicted the need for regular monitoring of the water bodies/wastewater as part of WBE studies in the urban and semi-urban areas to understand the outbreak and to assess spread of viral as well.

## Data Availability

All the data performed for this study is included in the preprint

## Authorship contribution statement

Manupati Hemalatha: Methodology, Investigation, Data curation, Writing – original draft. Athmakuri Tharak: Methodology, Investigation, Data curation, Writing – original draft. Harishankar Kopperi: Methodology, Investigation, Writing – original draft. Uday Kiran: Methodology, Investigation, Data curation, Writing – original draft. C.G. Gokulan: Methodology, Investigation, Formal analysis, Writing – original draft. Rakesh K. Mishra: Conceptualization, Supervision, Funding acquisition, Validation, Writing – review & editing. and S. Venkata Mohan: Conceptualization, Methodology, Supervision, Validation, Writing – original draft, Writing – review & editing.

## Declaration of competing interest

The authors declare that they have no known competing financial interests or personal relationships that could have appeared to influence the work reported in this paper.

## Acknowledgments

The work was supported by Council of Scientific and Industrial Research (CSIR), New Delhi, India in the form of project entitled ‘Testing for COVID-19 in wastewater as a community surveillance measure (6/1/COVID-19/2020/IMD)’. UK thanks UGC, CGG and MH thank CSIR for the financial support received. KH, AT, MH and SVM acknowledge the Director, CSIR-IICT for the support.

## Notes

### Competing Interest Statement

The authors have declared no competing interest.

### Author Declarations

The work was supported by Council of Scientific and Industrial Research (CSIR)New Delhi India in the form of project entitled Testing for COVID-19 in wastewater as a community surveillance measure (6/1/COVID-19/2020/IMD)

